# Understanding Adverse Population Sentiment Towards the Spread of COVID-19 in the United States

**DOI:** 10.1101/2021.07.15.21260543

**Authors:** Alexander Hohl, Moongi Choi, Richard Medina, Neng Wan, Ming Wen

## Abstract

**Background:** During the ongoing COVID-19 pandemic, the immediate threat of illness and mortality is not the only concern. In the United States, COVID-19 is not only causing physical suffering to patients, but also great levels of adverse sentiment (e.g., fear, panic, anxiety) among the public. Such secondary threats can be anticipated and explained through sentiment analysis of social media, such as Twitter.

**Methods:** We obtained a dataset of geotagged tweets on the topic of COVID-19 in the contiguous United States during the period of 11/1/2019 - 9/15/2020. We classified each tweet into “adverse” and “non-adverse” using the NRC Emotion Lexicon and tallied up the counts for each category per county per day. We utilized the space-time scan statistic to find clusters and a three-stage regression approach to identify socioeconomic and demographic correlates of adverse sentiment.

**Results:** We identified substantial spatiotemporal variation in adverse sentiment in our study area/period. After an initial period of low-level adverse sentiment (11/1/2019 - 1/15/2020), we observed a steep increase and subsequent fluctuation at a higher level (1/16/2020 - 9/15/2020). The number of daily tweets was low initially (11/1/2019 - 1/22/2020), followed by spikes and subsequent decreases until the end of the study period. The space-time scan statistic identified 12 clusters of adverse sentiment of varying size, location, and strength. Clusters were generally active during the time period of late March to May/June 2020. Increased adverse sentiment was associated with decreased racial/ethnic heterogeneity, decreased rurality, higher vulnerability in terms of minority status and language, and housing type and transportation.

**Conclusions:** We utilized a dataset of geotagged tweets to identify the spatiotemporal patterns and the spatial correlates of adverse population sentiment during the first two waves of the COVID-19 pandemic in the United States. The characteristics of areas with high adverse sentiment may be relevant for communication of containment measures. The combination of spatial clustering and regression can be beneficial for understanding of the ramifications of COVID-19, as well as disease outbreaks in general.

## Background

As of July 2021, The COVID-19 pandemic has caused over 184,000,000 confirmed cases and over 3,981,000 confirmed deaths globally since its onset in December 2019. In the United States (U.S.), over 33,000,000 cases were identified and over 605,000 deaths counted [1, 2, 3]. While vaccines have been developed and rolled out, various variants of the SARS-CoV-2 virus continue to pose a threat to public health [4, 5]. Strategies to slow the spread have been successful at reducing cases and deaths, but may have negative consequences on mental health, the economy, and society in general [6, 7, 8].

Humans under duress from constraints, such as fear, lack of resources, and quarantine, can behave in irrational, and sometimes violent ways [9, 10]. These behaviors include intensified racial hate, hate crimes, and hate-based violence [11, 12]. Therefore, the immediate threat of falling ill from COVID-19 is not the only concern. In the U.S., the increasing spread of COVID-19 is not only causing physical suffering to patients and an increasing burden to the healthcare system, but also great levels of adverse sentiment (e.g., fear, panic, anxiety) among the public [13]. Such secondary threats can be anticipated and explained through sentiment analysis of social media, such as Twitter [14].

Sentiment analysis (S.A.) “is the field of study that analyzes people’s opinions, sentiments, appraisals, attitudes, and emotions toward entities and their attributes expressed in written text.” [15]. Much S.A. research focuses on classifying written text, therefore it is considered part of the field of Natural Language Processing (NLP), even though it has been studied within the fields of Data Mining and Information Retrieval as well [15]. Applications of S.A. toward the COVID-19 pandemic include examinations of attitudes towards lockdowns [16], communication strategies of public health authorities [17], impacts of COVID-19 on mental health [18], and opinions towards vaccines [19], among others. The analysis of emotions is considered a subfield of S.A., as it focuses on revealing the psychological state of mind, e.g. of the author of a text document [20]. Emotions explicitly contain subjectivity, which distinguishes them from sentiments [21]. Words in context can be associated with basic emotions, such as *joy, sadness, anger, fear, disgust, surprise, trust*, and *anticipation* [22, 23]. This allows for classifying text documents using word-emotion dictionaries, such as the NRC Emotion Lexicon [24].

Understanding the geographic and temporal patterns of public sentiment, as well as its socio-demographic correlates, amid the COVID-19 pandemic, may allow public administrators to identify potential areas for proactive measures to prevent and mitigate social problems, as well as to optimize message targeting, government assistance, and overall communication strategies [25]. However, traditional survey or questionnaire-based data collection for assessing public sentiment may be difficult to implement in a timely manner during the pandemic. Instead, social media data such as geo-tagged tweets provide a feasible way to gain such information in near real-time [26, 27]. Social media usage has increased manyfold among American adults since 2005 [28], leading to a volume of 500 million tweets per day in 2017 [29]. Twitter has become an established data source in geographic research with applications in epidemiology [30], mobility pattern analysis [31], and disaster management [32], among others.

In this study, we use a curated dataset of geotagged tweets to analyze the spatiotemporal distribution of adverse sentiment toward the rapid spread of COVID-19 in the contiguous U.S. We identify spatiotemporal clusters of elevated adverse sentiment and present results in graphic and tabular form. In addition, we analyze the spatial correlates of adverse sentiment using a three-stage regression approach. This paper is structured as follows: The *Methods* section elaborates on the methods for *tweet data* collection, *sentiment classification, spatiotemporal clusters*, and the *spatial correlates of adverse sentiment*. This is followed by the *Results* section, which has two sub-sections that present the *spatial and spatiotemporal distribution* and the *spatial correlates of adverse sentiment*, respectively. Lastly, the *Discussion* and *Conclusions* sections contain a summary, assumptions and weaknesses, as well as relevance of this study within a broader context.

## Methods

### Twitter data

We obtained 6,080,400 tweets, which fulfilled the following selection criteria: 1) written in English, 2) geotagged, 3) sent from the contiguous United States, 4) sent between 11/1/2019 and 9/15/2020, 5) presence of COVID-19-related keywords (Table A1). Because this was a purchased dataset from Twitter, we were provided the entire set of requested tweets, rather than a sample, as provided as a free service through connection to the Twitter API. Geotagged tweets provide the user location if location services are enabled on the device. Depending on the quality of the global positioning system (GPS) signal, the geographic location of tweets may be exact or approximated to either cities, states, or nations. 83.67% of our tweets were geolocated at the city level, fulfilling the desired spatial granularity level of our analyses, and therefore retained for further processing. We then conducted a series of preprocessing steps to the tweet body: 1) removal of URLs and emojis, 2) removal of punctuation and other symbols (e.g. “@”, “&”), and 3) conversion of all letters to lower case.

### Sentiment classification

We classified our tweets using the word-emotion classification lexicon of the National Research Council Canada (NRC) [24]. The NRC Emotion Lexicon scores documents according to 8 different emotions (*joy, sadness, anger, fear, trust, disgust, surprise, anticipation*), based on the presence of emotional terms. For instance, the word “abandoned” is associated with the emotions of anger, fear, and sadness. Following the methodology of Karmegam & Mappillairaju [33], we considered the emotions of *sadness, anger, fear* and *disgust* as negative, and coded tweets that contained at least one of these emotions as “adverse”, whereas the remaining tweets are “non-adverse”. We used R for all data processing [34, 35], and specifically the “syuzhet” library to implement tweet classification [36]. In addition, we obtained the 2018 county polygon geometries as TIGER/Line shapefiles from the United States Census Bureau and used the “sf” library [37] to aggregate adverse and non-adverse tweets for each county within the contiguous United States (N = 3108), for each day within our study period (N = 238).

### Spatiotemporal clusters of adverse sentiment

Using the county-level, daily counts of adverse and non-adverse tweets, we employ the retrospective space-time scan statistic [38] with Bernoulli model [39] to find clusters of adverse sentiment. This method identifies the most likely circular clusters out of a set of candidate clusters (a.k.a. “windows”), which are centered on the county centroids and defined by varying spatial and temporal search distances. We restricted the clusters spatially to include a maximum of 5% of all tweets, and temporally to last no longer than half of the study period.

Parameterization has a substantial impact on the size of the resulting clusters, as less restrictive parameter choices typically lead to larger clusters [40], necessitating careful optimization [41]. Our parameter choices are geared towards policymakers at the state-level, where the decisions on pandemic response are made [42], and therefore result in medium-sized clusters. The space-time scan statistic with Bernoulli model is used in a case-control scenario, such as adverse tweets vs. non-adverse tweets and employs a likelihood ratio test for each candidate cluster. The likelihood function is given in Equation 1 [43]:

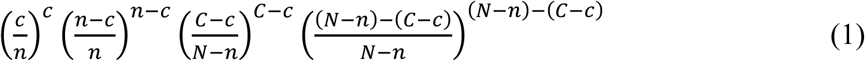

Where *C* is the total number of adverse tweets, *c* is the number of adverse tweets within the current candidate cluster, *N* is the total number of tweets within the entire study area/period, and *n* is the total number of tweets within the candidate cluster. The candidate with the highest value of the likelihood function is the strongest cluster, as it is a measure of risk within vs outside the cluster. We assess cluster significance using 999 Monte Carlo simulation runs, where tweet counts are randomized among the counties and days. Following the approach of [44, 45, 46], we map clusters together with the county-level proportion of adverse tweets. In addition, we report the following characteristics of statistically significant clusters at the *p* < 0.05 level: 1) the duration (*start* and *end* date), the number of counties contained within the cluster (# of *counties*); the relative risk (*RR*), which, similar to the likelihood function, is the ratio of the probability of observing an adverse tweet inside the cluster vs. outside; the observed number of adverse tweets (*Obs*); the expected number of adverse tweets (*Exp*) under the assumption that the number of adverse tweets (cases) follows the number of non-adverse tweets (controls); and the total number of tweets observed in the cluster (*Total tweets*).

### Spatial correlates of adverse sentiment

To understand the drivers of adverse population sentiment, we collected 9 county-level predictor variables from various sources (Table 1). The variables allow for a place-based description of counties in terms of the socioeconomic and demographic characteristics of the population. In addition, our predictor variables cover topics such as the rural-urban continuum, political views, social vulnerability, access to health care, and the severity and timing of the Covid-19 pandemic.

**Table 1.** Predictor variables.

| Variable | Source | Description |
| --- | --- | --- |
| *RUCC* | [47] | Rural-Urban Continuum Codes |
| *Ethnic* | [48] 2018 5-year ACS Estimates | Ethnic Heterogeneity Index |
| *Biden* | [50] | % Biden votes 2020 |
| *SVI 2* | [51] | Social Vulnerability Index Theme 2 |
| *SVI 3* |  | Social Vulnerability Index Theme 3 |
| *SVI 4* |  | Social Vulnerability Index Theme 4 |
| *PCP* | [52] | # primary care physicians per person |
| *Case rate* | [3] | COVID-19 cases per 100,000 people |
| *1<sup>st</sup> death* |  | # of days since 1<sup>st</sup> death |

The variables include: 1) The Rural-Urban Continuum Codes (*RUCC)* [47], which distinguish counties according to their population and proximity to metro areas. We reclassified the original 9 code values into metropolitan and non-metropolitan counties. 2) The Racial/Ethnic Heterogeneity Index (*Ethnic*) using the methodology by [48]:

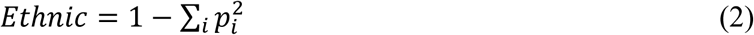

Where *p*_*i*_ is the proportion of the county population of the given group. For calculation of the index, we included the racial and ethnic categories: Latino/Hispanic, American Indian and Alaska Native, Asian, Black or African American, Native Hawaiian or Pacific Islander, and Non-Hispanic White [49]. The index ranges between 0-1, whereas a value of 1 indicates maximum heterogeneity. Conversely, a value of 0 indicates the presence of only one group. 3) The proportion of votes for Joseph R. Biden Jr. (*Biden*) in the 2020 election [50] as an indication of political views. The pandemic and its outcomes have been grossly politicized, and we suspect that political views have a substantial effect on adverse sentiment expressed in social media. 4) The 2018 Social Vulnerability Index (*SVI*) [51], which quantifies social vulnerability to natural hazards or disease epidemics. The SVI ranks counties according to four themes, which we use as variables in our modelling efforts: 1) socioeconomic status, 2) household composition, 3) race/ethnicity/language, and 4) housing/transportation. 5) The number of primary care physicians per county (*PCP*), which approximates access to healthcare [52]. The ease of receiving care may influence people’s feelings about the pandemic. Lastly, 6) The daily number of COVID-19 cases per 100,000 people (*Case rate*) using data from the John’s Hopkins Center for Systems Engineering [3]. As the temporal trend of the virus differs across counties, we took the day of the first observed death and calculated the number of days passed since the beginning of the study period (*1*^*st*^ *death*). The COVID-19-related variables are dynamic, as they vary through space and time, whereas the remaining variables are static, only varying through space.

To establish the correlates of adverse sentiment, we used ordinary least squares (OLS) regression, with the proportion of adverse tweets among all tweets as the dependent variable. We employed time flattening of our dynamic variables, including the dependent variable. This means for each county, we calculated the average of the time-series of our variables, effectively reducing a spatiotemporal dataset to a purely spatial one. We computed the variable correlation matrix to circumvent multicollinearity. Lastly, we ensured that variable inflation factor (VIF) values of our predictor variables remained below an acceptable level of 2.5 [53]. We checked for heteroskedasticity using residual vs. fitted values and Q-Q plots. Further, we checked for spatial autocorrelation of model residuals using Moran’s I [54]. Since our data is highly zero-inflated (many rural counties have zero adverse tweets), we ran zero-inflated beta regression [55] with the variables we determined through OLS.

Due to the locational uncertainty associated with geotagged tweets, and to address zero-inflation of the county-level tweet counts, we built an additional model with a smoothed response variable using kernel density estimation, as in Equation 3:

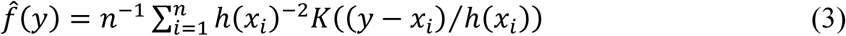

Here, *x*_*i*_ refers to the *i*^*th*^ observation (county), *h(x)* is the bandwidth function, and *K(*.) is the kernel function. We used adaptive bandwidths due to the variable spacing of county centroids across the United States (counties in the western United States are larger in general than counties in the east). The variable bandwidths are governed by specification of a smoothing multiplier (a.k.a “global bandwidth”), calculated using the oversmoothing principle [56]. We weighted the resulting densities by adverse/total tweet count separately, and computed the relative risk function, as in Equation 4:

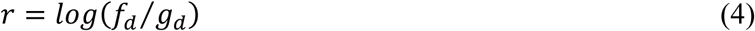

where *f*_*d*_ and *g*_*d*_ refer to the densities of adverse and total tweets, respectively. The log transformation allows for symmetric treatment of the two densities [57, 58]. Since the county centroids are the geographic representation of all our observations, we extracted the resulting risk values at their locations. We employed the kernel density routines implemented in the “sparr” library in R [59, 60, 61].

## Results

### Spatial and temporal distribution of adverse sentiment

Adverse sentiment expressed in tweets varies substantially throughout our study period (Figure 1). The temporal pattern of adverse sentiment is characterized by an initial period of fluctuation at a low level (∼30% - 37%) from 11/1/2019 - 1/15/2020, followed by a steep increase and subsequent fluctuation at a higher level (∼40% - 48%) from 1/16/2020 - 9/15/2020. The total number of daily tweets remained around zero from 11/1/2019 - 1/22/2020, followed by an increase to up to 80,000 daily tweets around 3/15/2020. The number of tweets subsequently decreases and remains relatively stable at around 10,000 – 15,000 until the end of the study period.

**Figure 1.**
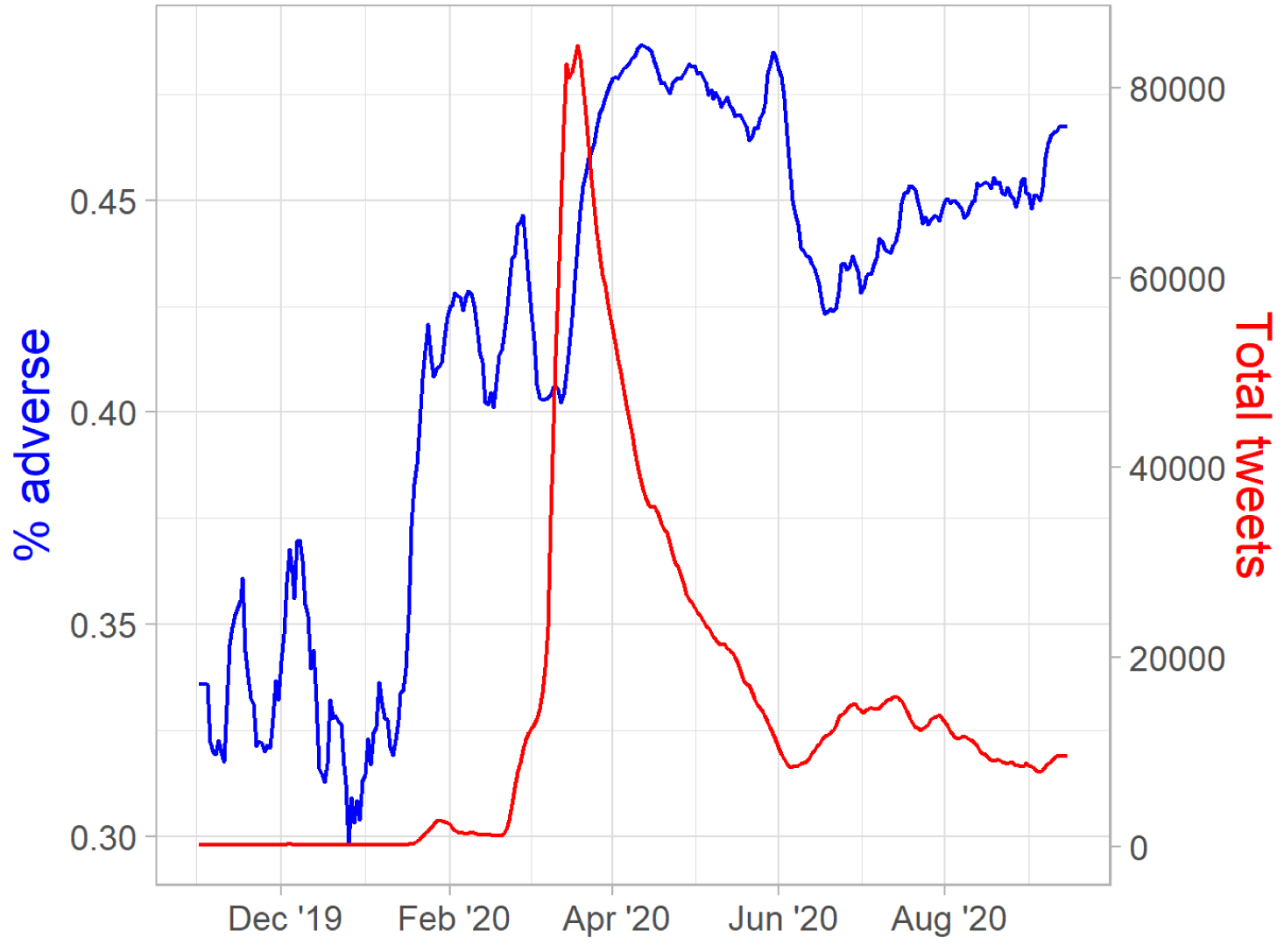
Temporal distribution of total tweets (red line) and the proportion of adverse tweets (blue line).

Using the retrospective space-time scan statistic with Bernoulli model, we identified 12 clusters of elevated adverse sentiment, as expressed in tweets (Figure 2, Table 2). The clusters are spread across the entire contiguous U.S., except for portions of the Midwest and Pacific Northwest regions. The clusters exhibit substantial variation in size, as their radii range from 765 km (Cluster 4) to 31 km (Cluster 2). All clusters are active during the distinct period from late March to May/June 2020. Cluster 1 includes 136 counties of California, Oregon, Idaho, Nevada, Wyoming, Utah, Colorado and Arizona and contains 69,842 adverse tweets (*Obs*, Table 1), whereas only 64,155 are expected (*Exp*). The total number of tweets for Cluster 1 is 141,413 (*Total tweets*), therefore the relative risk (*RR*) is elevated at 1.09. Cluster 2 is much smaller than Cluster 1 as it merely encompasses 4 counties of the New York City area. The observed and expected number of adverse tweets are lower (46,607 and 42,209, respectively), but the *RR* is higher (1.11). Similar to Cluster 2, Cluster 3 is small, as it consists of only 2 counties in Vermont. Expectedly, the *Total tweets* are low (2,667). However, it has an RR of 1.55, which is the highest value across all clusters. The largest cluster (Cluster 4) includes 237 counties in Texas, Arizona and New Mexico, and has an *RR* of 1.08. Cluster 5 consists of 111 counties in the lower Michigan peninsula, Ohio, Indiana and Illinois. It includes the cities of Chicago and Detroit and has the highest *Total tweets* among all clusters (148,807). Cluster 6 consists of 128 counties in New York, Pennsylvania, New Jersey, Virginia and Ohio, has an *RR* of 1.07 and is of medium size. Cluster 7 has a total of 421 counties, which is the highest number among all clusters, but an *RR* of 1.06, which puts it in the lower third. Cluster 8 consists of 4 counties in southern California which include the Los Angeles and San Diego metro areas. Cluster 9 is in the Appalachian Mountains around Georgia, the Carolinas, Tennessee, Kentucky and West Virginia. Encompassing 320 counties, it is the second highest in that category. Cluster 10 includes 53 counties surrounding the Chesapeake Bay in Maryland and Delaware. Cluster 11 covers the southern half of Florida, including large cities of Miami, Tampa, and Orlando. Finally, Cluster 12 includes 79 counties of Colorado, Kansas, Nebraska, South Dakota and Wyoming.

**Figure 2.**
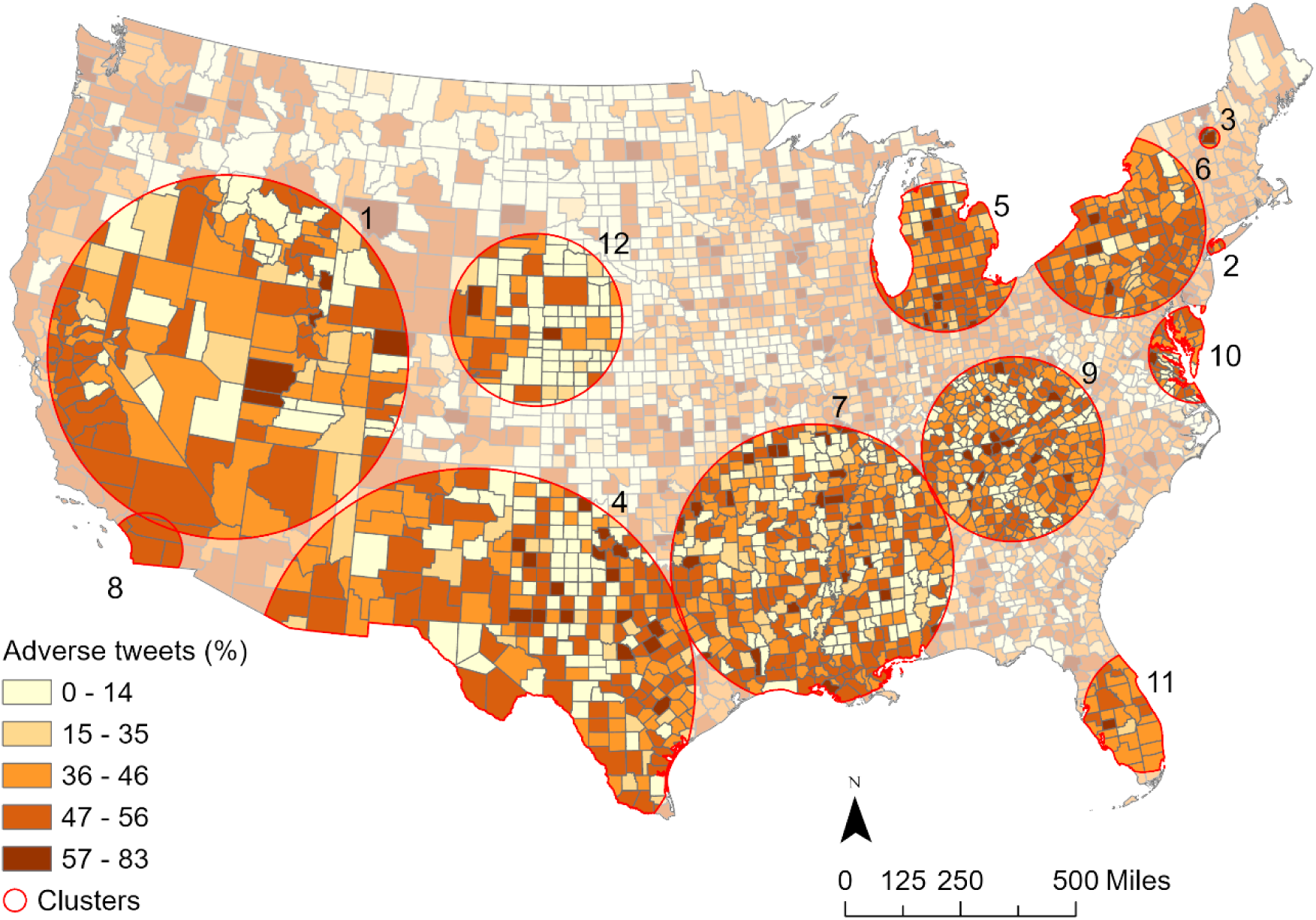
Spatial distribution of adverse tweets augmented by clusters resulting from the space-time scan statistic.

**Table 2.**
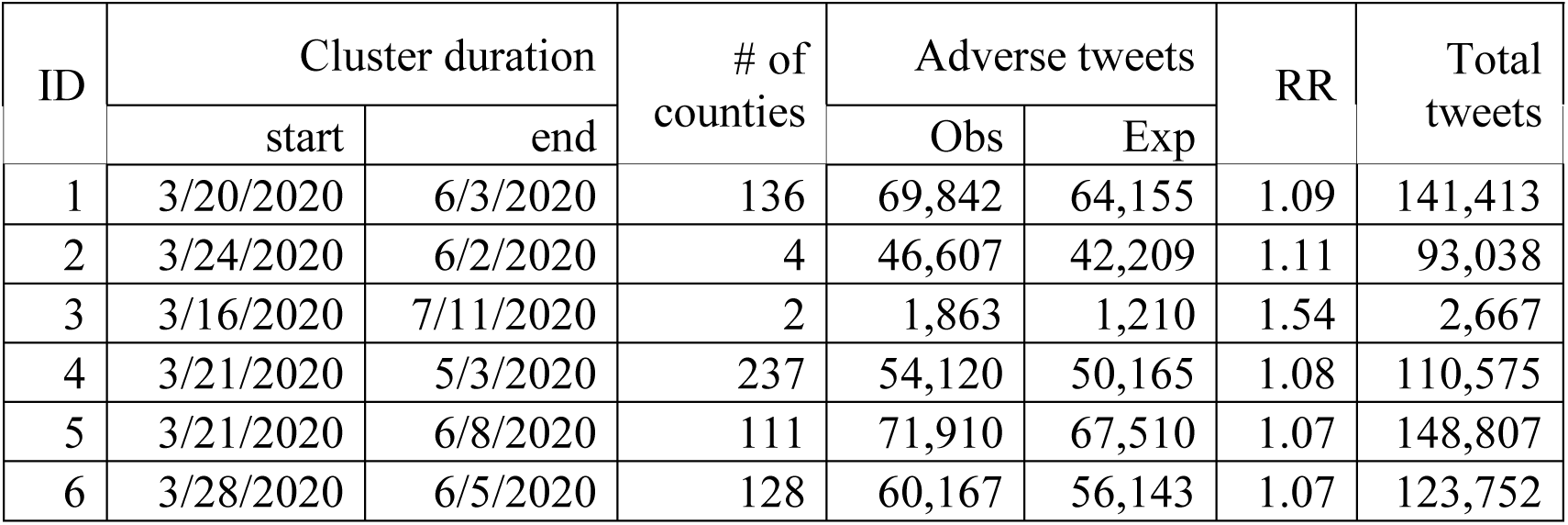
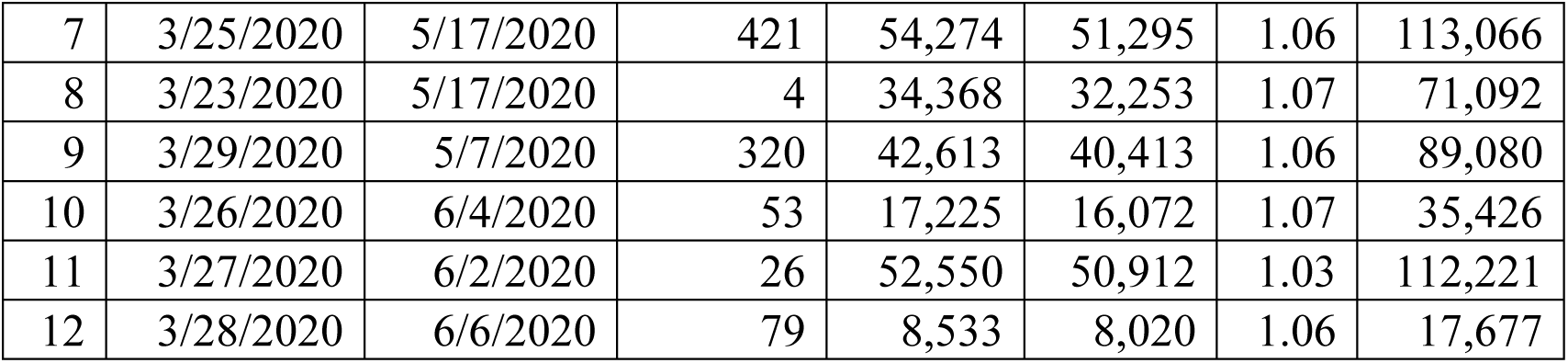
Clusters from the space-time scan statistic (corresponding to Figure 2).

### Spatial correlates of adverse sentiment

Figure 3 illustrates the spatial distributions of our predictor variables, which exhibit some patterns worth noting: the *Ethnic* variable exhibits high values in the coastal and southern regions of the United States, whereas the central and northern regions are more homogeneous. The *RUCC* variable traces metropolitan regions, which are predominantly found on the coasts, whereas rural counties cover most of the Midwest and West. Support for Joseph R. Biden Jr. in the 2020 election (*Biden*) mostly came from urban regions. Notable exceptions include the “black belt” in the Southeast [62] and rural counties in Arizona, Colorado and New Mexico.

**Figure 3.**
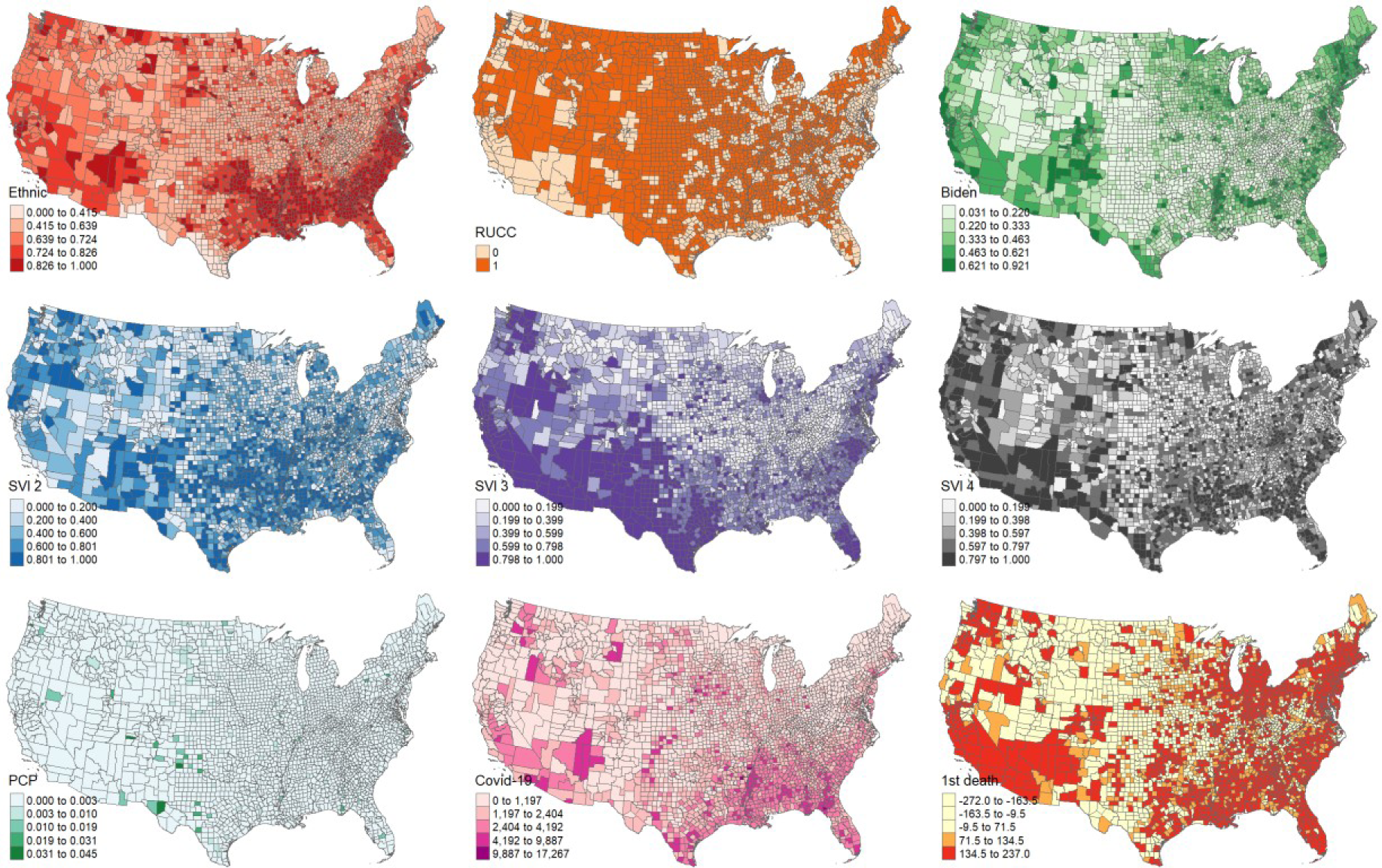
Predictor variable maps.

All chosen variables are significant in the multivariate OLS model (Table 3). It shows that higher adverse sentiment is associated with counties that are less heterogeneous (*Ethnic* variable), less rural (*RUCC1*), had higher support for Joseph R. Biden Jr. (*Biden*) in the 2020 election, are less vulnerable in terms of household composition and disability (*SVI 2*), but more vulnerable in terms of minority status and language (*SVI 3*), as well as more vulnerable in terms of housing type and transportation (*SVI 4*), have a lower number of primary care physicians (*PCP*), have a lower COVID-19 case rate (*Case rate*), and had their first death due to COVID-19 early on (*1*^*st*^ *death*). The adjusted R-squared of the OLS model is 0.593, which indicates a good model fit. The zero-inflated (zi) beta regression model largely echoes the findings of the OLS model, as all variables are significant as well, but the estimates are generally higher, indicating a stronger effect of the variables. The Akaike Information Criterion (AIC) for the OLS model is lower than for the zi beta regression model (−5711.43 vs. -1795.01), indicating a better fit of the zi model.

**Table 3.**
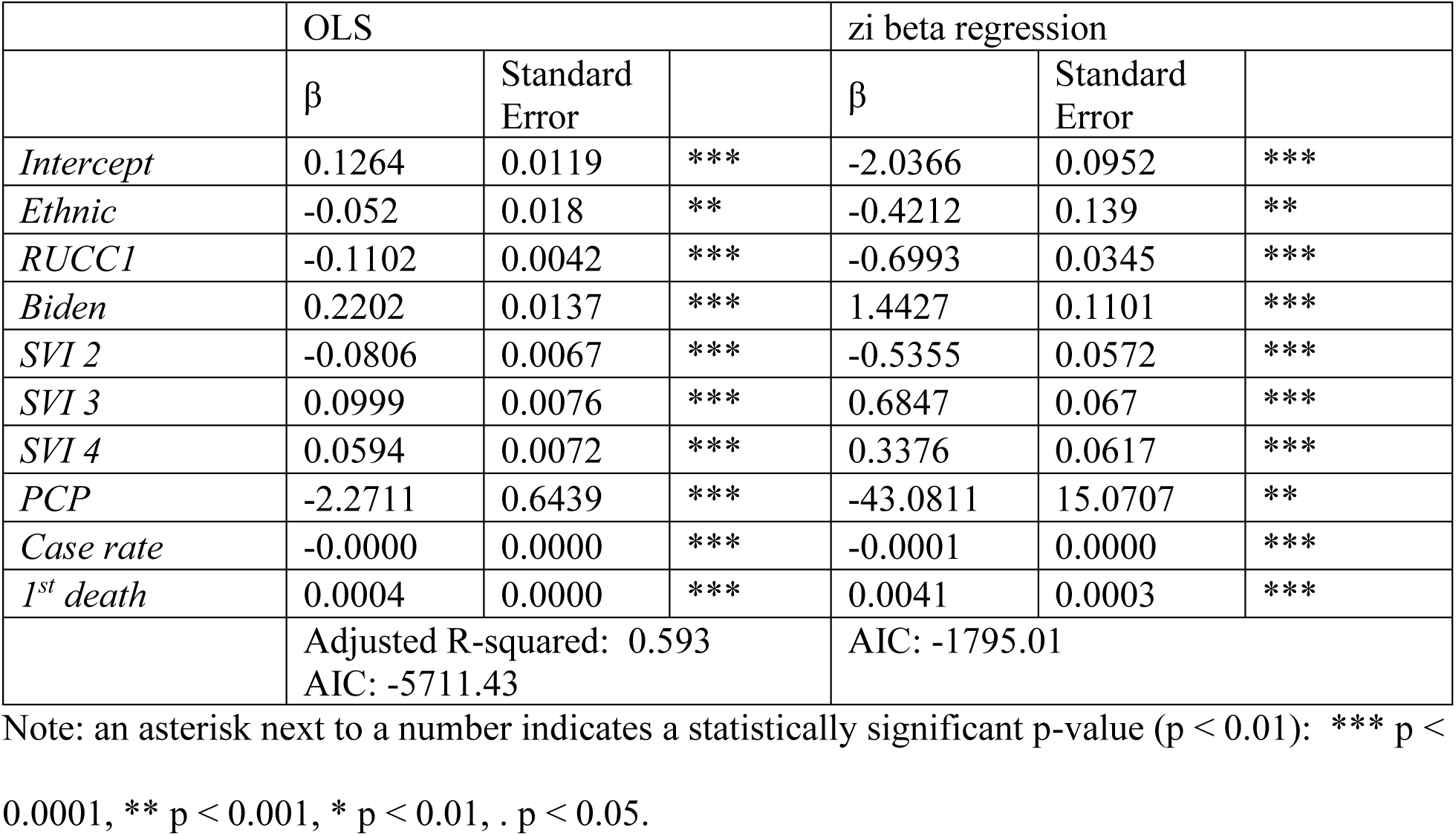
Multivariate models.

**Table 4.**
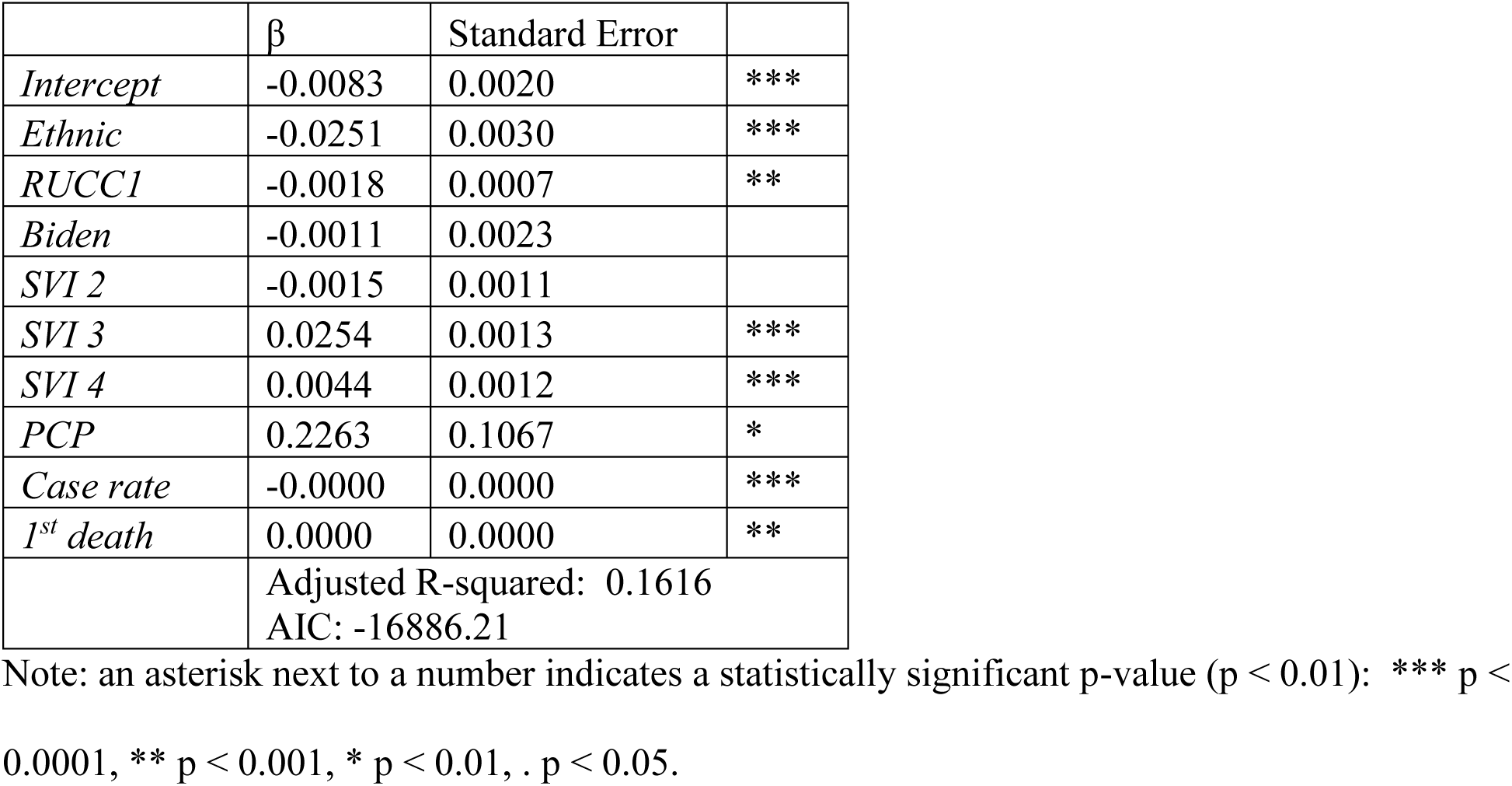
OLS model with smoothed response variable.

Both, the OLS model and the zi beta regression model suffer from trend in the residuals, as indicated in their residuals vs. fitted plots (Figure 4). The OLS model residuals exhibit a negative relationship with the fitted values (Figure 4 A), which indicates heteroskedasticity, a violation of OLS assumptions. The zi beta regression model residuals (Figure 4 C) show a decreasing variance with increasing fitted values, also indicating heteroskedasticity. Q-Q plots of both, the OLS model (Figure 4 B) and the zi beta regression model (Figure 4 D), show a deviation from normality, especially towards the tails of the distribution. In summary, the diagnostics of both, the OLS and the zi beta regression models show violations of regression assumptions, hence the validity of the model coefficients presented in Table 3 is questionable.

**Figure 4.**
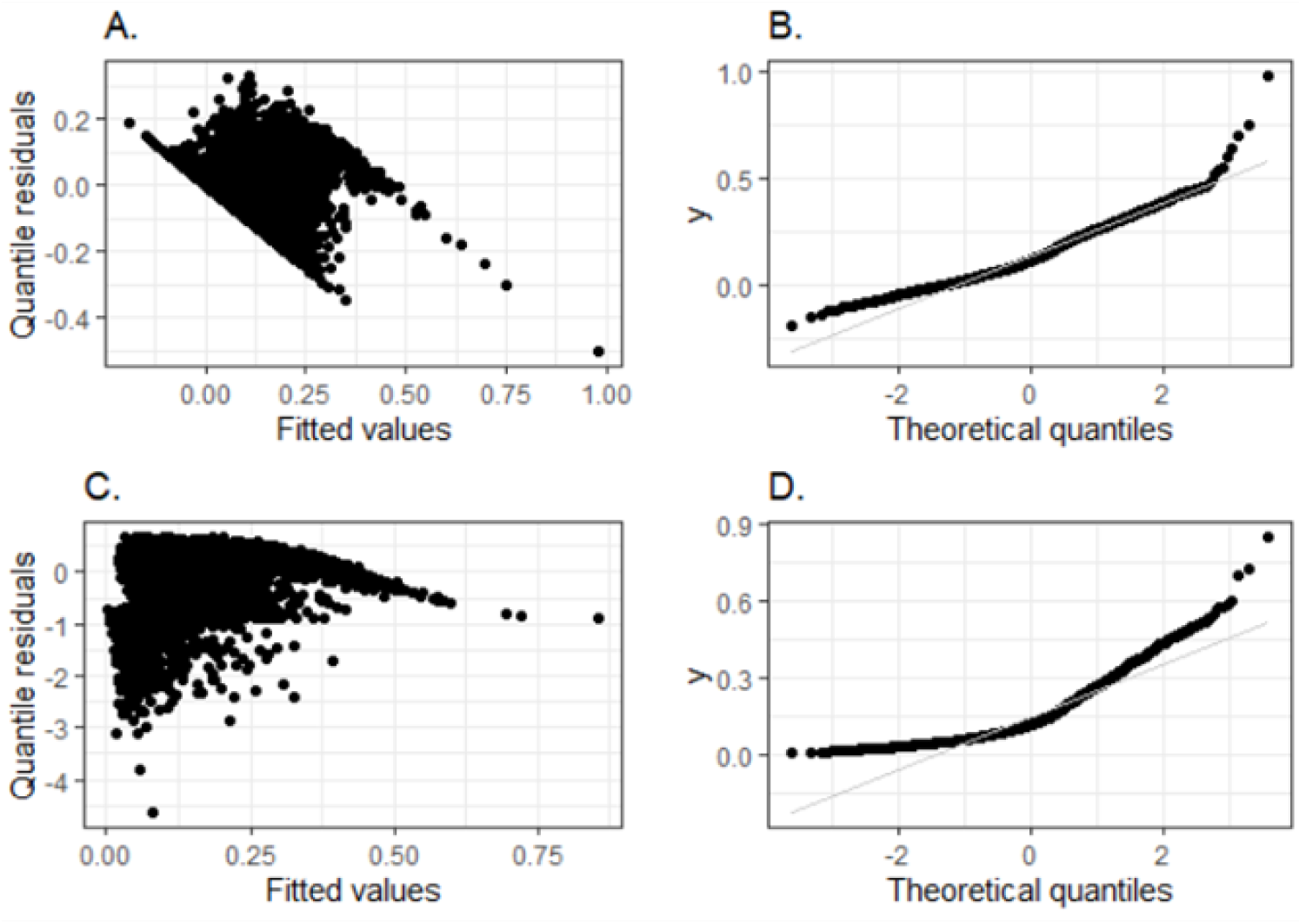
Model diagnostics. A. Residuals vs. Fitted plot of OLS model, B. Q-Q plot of OLS model, C. Residuals vs. Fitted plot of zi beta regression model, D. Q-Q plot of zi beta regression model.

The smoothed response variable shows variation in risk (proportion of adverse tweets) across the study area (Figure 5). It is apparent that major urban areas like New York City, Chicago, Los Angeles and San Francisco are clusters of adverse sentiment. Conversely, rural areas in the Midwest, the Carolinas and in Maine show little adverse sentiment.

**Figure 5.**
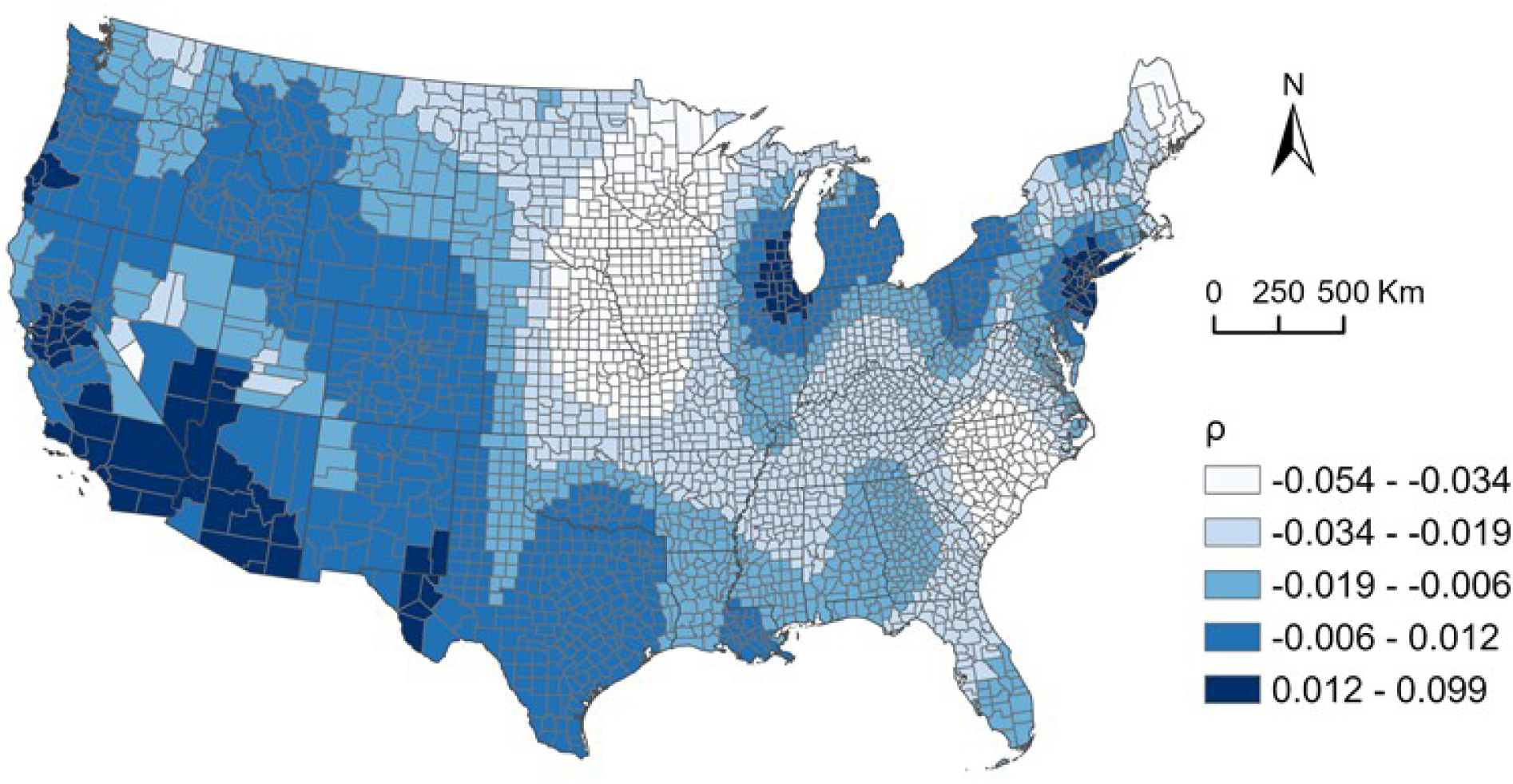
Smoothed response variable.

The OLS model with smoothed response variable shows some differences to the previous two models: The *Biden* and *SVI 2* variables are not significant, indicating that political views and vulnerability in terms of household composition and disability do not have an effect on adverse sentiment. In addition, the *PCP* variable has reversed it sign from negative to positive compared to the previous two models. It means that counties with a higher number of primary care physicians observe a higher share of tweets classified as “adverse”. However, its standard error is quite high (0.1067) and the variable is only significant at the p < 0.01-level and would not pass a more rigorous significance test. The adjusted R-squared (0.1616) indicates a lower model fit, but with an AIC of -16886.21, it is preferable over the previous two models due to relatively less information loss.

Model diagnostics also show an improvement of the smoothed response variable over the previous models, as there is no apparent trend recognizable in the residuals vs. fitted plot (Figure 6 A), and while residuals still deviate from normalcy, these deviations are found at the tails of the distribution. Because the Moran’s I of model residuals was 0.02 (p=0.04), which means barely significant light clustering, we omitted further investigation into spatial regression. In summary, the three models allow us to draw a valid picture of the spatial correlates of adverse sentiment.

**Figure 6.**
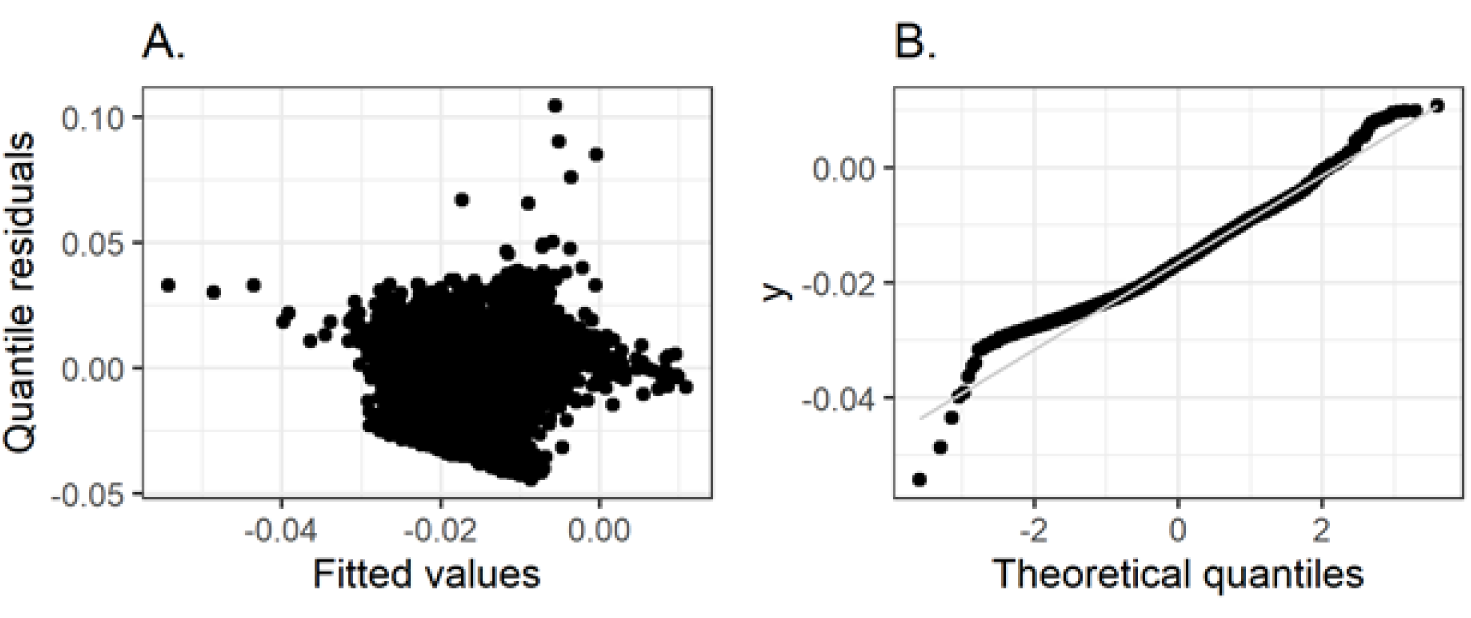
Model diagnostics: OLS model with smoothed response variable. A. Residuals vs. Fitted plot, B. Q-Q plot.

## Discussion

In this study, we analyzed a dataset of geotagged tweets with focus on adverse sentiment towards the spread of COVID-19. We identified spatiotemporal clusters of adverse sentiment in the contiguous U.S. at the county-level. In addition, we employed sentiment analysis to find the spatiotemporal distribution of adverse sentiment and identified spatial correlates using regression models. Our findings indicated that adverse sentiment is increased in counties that are racially/ethnically homogeneous, urban, vulnerable due to minority status and language, as well as housing type and transportation, have lower COVID-19 prevalence rates, and experienced mortality due to COVID-19 at an early stage of the pandemic. To our knowledge, this is the first study that uses sentiment analysis, clustering methods, and regression modeling to identify areas of adverse sentiment, and to identify associations of such sentiment with area socioeconomic and demographic characteristics.

Using the space-time scan statistic is attractive because it identifies four important cluster characteristics: 1) geographic location and extent, 2) temporal duration, 3) statistical significance, and 4) strength (relative risk). It retrospectively identifies areas of concern, which allows for evaluating policy to slow the spread of COVID-19, as well as communication strategies of health authorities. The clusters we identified cover the entire spectrum of urban to rural areas. However, for some urban areas, the central county is not part of the cluster, while its surrounding suburban areas are. Examples include New York City, where New York County (Manhattan) is not within the cluster, but the neighboring counties are (Kings, Queens and Bronx and Nassau Counties). We observe the same pattern for Washington D.C., San Francisco, and Los Angeles. This points towards a disparity between central cities and their suburbs, which health authorities should be aware of.

Our regression modelling approach allowed us to confirm robust relationships between adverse sentiment and various socioeconomic and demographic predictor variables. Some of the results seem counterintuitive at first: 1) the negative association of rurality and adverse sentiment was not expected. However, most tweets are sent from urban/suburban places, therefore they might not be as representative for population sentiment in rural areas. 2) The coefficients for the COVID-19 case rate are significant in our models, but too small to distinguish from zero, given the precision of our reporting. For instance, the coefficient for the COVID-19 case rate in the zero-inflated beta regression model is -0.0001, which means that adding 100,000 cases leads to a 0.0001 decrease in the proportion of adverse tweets, a very small difference. Therefore, even though the coefficient is significant, we do not think this is a meaningful relationship. The same can be said about the timing of the first death, where adding one day leads to an increase of adverse tweets of 0.0041. 3) Support for Joseph R. Biden Jr. was not significantly associated with adverse sentiment in the final OLS model with smoothed response variable. This can possibly be explained by our sole focus on adverse sentiment, and not the subject of the adverse sentiment: people on both sides of the political spectrum may express their frustration and anger.

Despite the merits of our study, we want to point out the following weaknesses and future research directions: 1) While a major strength of the lexicon-based sentiment classification approach lies in its simplicity, this method does not allow us to identify more complicated language features, such as sentiment shifters (e.g., “I don’t like this car”, is negative, even though the word “like” is not; [15]); 2) Twitter users represent a younger demographic group, whose sentiments and opinions may not reflect those of the entire population. In addition, urban areas tend to be overrepresented in tweet samples [63]. We tried to partially address this issue by including the proportion of the population between 18 and 34 (the main demographic who uses Twitter) but discarded the variable during our modelling process due to unacceptably high correlations with other variables on our model. 3) Our regression modelling approach does not consider the temporal dimension (except for the *1*^*st*^ *death* variable), despite having a spatiotemporally complete dataset. Therefore, our current and future research efforts focus on the application of spatiotemporally explicit modelling using Bayesian statistics to address the spatial and temporal nature of our dataset [64]. Lastly, due to the real-time availability of data, such as tweets and various metrics on COVID-19, it is feasible to apply our methods and update the results of this study daily. For instance, the space-time scan statistic can be employed in a prospective fashion, which identifies current and emerging clusters exclusively [44, 45]. Similarly, the regression models can be updated as new data becomes available.

## Conclusions

We utilized a dataset of geotagged tweets to identify the spatiotemporal patterns and the spatial correlates of adverse population sentiment during the first two waves of the COVID-19 pandemic in the United States. We recommend that disease response in counties belonging to clusters of adverse sentiment needs to consider the characteristics we identified in our regression modelling approach. The combination of spatial clustering and regression can be beneficial for understanding of the ramifications of COVID-19, as well as disease outbreaks in general. Except for the original tweets, all our data, software and code are publicly available through a GitHub repository^1^. The dissemination of reproducible research is crucial, as it allows other scientists to contribute to our knowledge about the COVID-19 pandemic. Geographers can play a vital role in understanding disease outbreaks, as well as their consequences for society. This study is one example out of many methods that can be applied in limited time to guide public health authorities in their response to spatial and spatiotemporal disease transmission dynamics.

## Data Availability

The datasets generated and/or analyzed during the current study are available in the covid19sentiment repository, https://github.com/alexandster/covid19sentiment

## Abbreviations

U.S.: United States of America
S.A.: Sentiment Analysis

## Declarations

### Availability of data and materials

The datasets generated and/or analyzed during the current study are available in the covid19sentiment repository, [hidden for peer review]

### Authors’ contributions

**AH** conceived the study, drafted the manuscript and performed statistical analyses. **MC** performed data wrangling. **RM** conceived the study, acquired the data, and drafted the manuscript. **NW** conceived the study and edited the manuscript. **MW** conceived the study, edited the manuscript and contributed expertise on data analysis.

## Acknowledgements

The authors acknowledge Dr. Simon Brewer (University of Utah), who contributed expertise on spatiotemporal statistics.

## Supplementary Information

Additional file: Table A1

## Footnotes

1 [hidden for peer-review]

