## Supplemental File 1 for "Understanding Adverse Population Sentiment Towards the Spread of COVID-19 in the United States"

Table A1. COVID-19 ruleset.

Ruleset: (#Coronavirusmexico OR #covid2019 OR #coronavirususa OR #covid_19uk OR #covid-19uk OR #Briefing_COVID19 OR #coronaapocolypse OR #coronavirusbrazil OR #marchapelocorona OR #coronavirusbrasil OR #coronaday OR #coronafest OR #coronavirusu OR #covid2019pt OR #COVID19PT OR #caronavirususa OR  covid19india OR #caronavirusindia OR #caronavirusoutbreak OR #caronavirus OR carona virus OR #2019nCoV OR 2019nCoV OR #codvid_19 OR #codvid19 OR #conronaviruspandemic OR #corona OR corona OR corona vairus OR corona virus OR #coronadeutschland OR #Coronaferien OR #coronaflu OR #coronaoutbreak OR #coronapandemic OR #Coronapanik OR #cronapocalypse OR #CoronaSchlager OR #coronavid19 OR #coronavid19 OR #Coronavirus OR Coronavirus OR #coronavirusargentina OR #coronavirusbrasil OR #CoronaVirusCanada OR #coronaviruschile OR #coronaviruscolombia OR #CoronaVirusDE OR #coronavirusecuador OR #CoronavirusEnColombia OR #coronavirusespana OR CoronavirusFR OR #CoronavirusFR OR #coronavirusIndonesia OR #Coronavirusireland OR #CoronaVirusIreland OR #coronavirusmadrid OR #coronavirusmexico OR #coronavirusnobrasil OR #coronavirusnyc OR #coronavirusoutbreak OR #coronavirusoutbreak OR #coronaviruspandemic OR #coronavirusperu OR #coronaviruspuertorico OR #coronavirusrd OR #coronavirustruth OR #coronavirusuk OR coronavirusupdate OR #coronavirusupdates OR #coronavirusuruguay OR "coronga virus" OR corongavirus OR #Corvid19virus OR #covd19 OR #covid OR covid OR #covid OR covid OR covid 19 OR #covid_19 OR #covid_19 OR Covid_19 OR #COVID_19uk OR #covid19 OR Covid19 OR Covid19_DE OR #covid19Canada OR Covid19DE OR Covid19Deutschland OR #covid19espana OR #covid19france OR #covid19Indonesia OR

#covid19ireland OR #covid19uk OR #covid19usa OR #covid2019) lang:en has:geo place_country:US

(#ForcaCoronaVirus OR #infocoronavirus OR #kamitidaktakutviruscorona OR #nCoV OR nCoV OR #ncov2019 OR nCoV2019 OR NeuerCoronavirus OR #NeuerCoronavirus OR Nouveau coronavirus OR #NouveauCoronavirus OR "novel coronavirus" OR #NovelCorona OR novelcoronavirus OR #novelcoronavirus OR #NovelCoronavirus OR #NuovoCoronavirus OR #ohiocoronavirus OR #PánicoPorCoranovirus OR #SARSCoV2 OR #SARSCoV2 OR "the coronas" OR #thecoronas OR #trumpdemic OR "Virus Corona" OR #viruscorona OR #CoronaAlert OR #coronavirusUP OR #coronavirustelangana OR #coronaviruskerala OR #coronavirusmumbai OR #coronavirusdelhi OR #coronavirusmaharashtra OR #coronavirusinindia OR #covid_19ind OR #covid19india OR "coronavirus india" OR #coronavirusindia OR #bayarealockdown OR #stayathomechallenge OR #stayhomechallenge OR #quarantinelife OR #dontbeaspreader OR #stayhomechallenge OR #howtokeeppeoplehome OR #togetherathome OR "alcool em gel" OR "alcool gel" OR

#alcoolemgel OR #alcoolgel OR #avoidcrowds OR "bares cerrados" OR "bares fechados" OR "bars closed" OR #canceleverything OR #CerradMadridYa OR "clases anuladas" OR #CLOSENYCPUBLICSCHOOLS OR #confinementtotal OR #CONVID19 OR #CoronavirusESP OR cuarentena OR #cuarentena OR #CuarentenaCoronavirus OR #cuarentenaYA OR "dont touch ur face" OR "dont touch your face" OR #DontBeASpreader OR #donttouchyourface OR "escolas fechadas" OR "escolas fechando" OR "escolas sem aula" OR "escolas sem aulas" OR #euficoemcasa OR "evitar el contagio" OR #ficaemcasa OR "flatten the curve" OR "flattening the curve" OR #flatteningthecurve OR #flattenthecurve OR #FrenaLaCurva OR "Hand sanitizer" OR #Handsanitizer OR #HoldTheVirus OR "lava tu manos" OR #lavatumanos OR "lave as maos" OR #laveasmaos OR #lockdown OR lockdown OR #pandemic OR pandemic

OR #panicbuying OR #panickbuing OR quarantaine OR #quarantine) lang:en has:geo place_country:US

(quarantine OR #QuarantineAndChill OR quarantined OR quarentena OR #quarentena OR #quarentine OR #quarentined OR quarentined OR #quarentinelife OR quedateencasa OR #remotework OR #remoteworking OR "restaurantes cerrados" OR "restaurantes fechados" OR "restaurants closed" OR #selfisolating OR #SiMeContagioYo OR "social distancing" OR #socialdistance OR #socialdistancing OR #socialdistancingnow OR #socialdistnacing OR #stayathome OR #stayathome OR #stayhome OR #stayhome OR #stayhomechallenge OR #stayhomesavelives OR #StayTheFHome OR #StayTheFuckHome OR #suspendanlasclases OR teletrabajo OR #teletrabajo OR #ToiletPaperApocalypse OR #toiletpaperpanic OR "trabajadores a la calle" OR "trabajar desde casa" OR #trabajardesdecasa OR "trabalhando de casa" OR "trabalhar de casa" OR "wash ur hands" OR "wash your hands" OR #washurhands OR #washyourhands OR #WashYourHandsAgain OR #wfh OR "work from home" OR #workfromhome OR "working from home" OR #workingfromhome OR yomequedoencasa OR #2019_ncov OR #21dayslockdown OR #5baje5minute OR #9minute9baje OR #9minutesforindia OR #AislamientoObligatorio OR aksiberantasCovid19 OR #AlertaCOVID19SV OR #ampliarlacuarentenaes OR #auxilioemergencial OR #BersamaMelawanCorona OR #bersatulawancovid19 OR #bloqueioderuas OR #Bogotaencasa OR #bolsonarogenocida OR #BreakCorona OR #cadeostestes OR #californialockdown OR #californiaquarantine OR #californiashutdown OR #calockdown OR #capitaocorona OR #CegahTangkalCorona OR #chegadequarentena OR #clapforourcarers OR #ClubQuarantine OR #CoronaChainScare OR

#CoronaCitizenResponsibility OR #CoronaCrises OR #coronacrises OR #CoronaCrisis OR #coronafreepakistan OR #CoronaInPakistan OR #coronakrise OR #coronalockdown OR #coronanasperiferias OR #coronapandemie OR #coronapandemie OR #CoronaStopKaroNa OR #coronaupdate OR #coronaupdatesindia OR #CoronaVillains OR #coronaviru OR #CoronavirusBillUK OR #CoronaVirusFromMinorChastisement OR #coronavirusinsa OR #coronavirusitalia OR #coronavirusitaly) lang:en has:geo place_country:US (#coronaviruslombardia OR #coronavirusoubreak OR #coronavirusplantao OR #CoronavirusPlantao OR #CoronaVirusSA OR #CoronavirusSchweiz OR #coronavirussouthafrica OR #CoronavirusSuisse OR #CoronavirusSwitzerland OR #coronavoucher OR #coronawarriors OR #coronawarriors OR #coronazeit OR #coronovirus OR #coronvirus OR #corronavirus OR #Covid19_CH OR #covid19brasil OR #Covid19CH OR #covid19ecuador OR #Covid19InSA OR #COVID19italia OR #covid19out OR #Covid19Schweiz OR #covid19southafrica OR #covidactnow OR #covideos OR #covididiots OR #covidiots OR #covidsafe OR #CuarentenaHastaJunioEs OR #CuarentenaInformando OR #cuarentenametadata OR #cubaporlasalud OR #cubasalvavidas OR #CubriendoElCoronavirus OR #CurfewInIndia OR #CurfewInIndia OR #CuronaVairus OR #curonavirus OR #depoisdaquarentenaeu OR #dirumahaja OR #disciplinaparavolver OR #disiplincegahcorona OR #ecuadorencrisis OR #ecuadorenemergencia OR #eunaquarentena OR #FightCoronaWithJokowi OR #fiqueemcasa OR #FlexibilizarElAislamientoEs OR #frontlinewarriorsintern OR #frontlineheroes OR #frontlineworkers OR #frontlineworkersappreciation OR #GerakanSocialDistancing OR #homeschool OR #homeschooling OR #HomeTasking OR #hydroxychloroquine OR #indiafightcorona OR #indiaprotectdoctors OR #indonesialawancorona OR #indonesialawancovid19 OR #JagaDiriJagaJarak OR

#JanataCurfewMarch22 OR #JantaCurfewChallenge OR #KanikaCoronaRow OR #KanikaKaCoronaCrime OR #koronaindonesia OR #koronavirusIndonesia OR #LaPrevenciónEstáEnTusManos OR #LawanCorona OR #LawanCoronaBersama OR #lawancovid19 OR #LawanCovid19 OR #LetsDefeatCOVID19Together OR #Lockdowhustle OR #lockdown21 OR #LockdownEnd OR #LockdownNow OR #lockdowntillmay3 OR #losangeleslockdown OR #mascaras OR #mascarasalva OR #mascarilla OR #mascarillas OR #mascarillassolidarias OR #masks4all OR #maskuplagos OR #micasaesmiplaza OR #michiganshutdown) lang:en has:geo place_country:US

(#ModiKiBaatMano OR #mp927 OR #mpdafome OR #mpdamorte OR #mpdobolsonaro OR #mumbailockdown OR #n95 OR #NeuerCoronavirusSchweiz OR #NoAlAislamientoInteligente OR #NotDying4WallStreet OR #notessential OR #obrasilnaopodeparar OR #OBrasilnaoVaiParar OR #obrasilvaiparar OR #OneTeamFromHome OR #onlineclasses OR #ParemosElVirus OR #PMCares OR #PPE OR #ppeshortage OR #pralernaquarentena OR #PrioridadDineroOSalud OR #QuarantineMoneyMakingIdeas OR #quaratinelife OR #quarentenabrasil OR #quarentenaLGBTQ OR #quarentenou OR #quaretenabrasil OR #QuaronaVirus OR #ReceitasDaQuarentena OR #remdesivir OR #SCNaoQuerMorrer OR #selfemployedmattertoo OR #SendUsBackHome OR #ShamblesStayAtHome OR #sideeffectsofquarantinelife OR #Social_Distancing OR #sosecuador OR #spcontraocoronavirus OR #SRKDonatesForCovid OR #stayathomeorder OR #StayAtHomeReadABook OR #StayAtHomeSaveLives OR #stayhomestaysafe OR #suspendonlineclasses OR #taxarfortunassalvarvidas OR #TaxarFortunasSalvarVidas OR #TaxarFortunasSalvarVidas OR #testesmasivosja OR #testesmassivosja OR #ThaliBajao OR #thankyouwarriors OR #UKCoronavirusBill OR #uklockdown OR

#UNExigimosGarantiasLaborales OR #unidosvenceremos OR #UNSuspendanClasesOParamos OR #unsuspendanclasesya OR #UNSuspendanlasClasesOParamos OR #usemascara OR #vaipassar OR #vegasshutdown OR #viviremosyvenceremos OR #VivirEnCuarentenaEs OR #WarAgainstVirus OR #waragainstvirus OR #wearamask OR #WhenCoronaVirusIsOver OR #WhoPaysForCovid OR #WorkingFromHomeLife OR #workingfromhometips OR "aula online" OR "auxilio emergencial" OR "Bloody Diwali" OR "bloqueio de ruas" OR "bloqueio de vias" OR "bloqueio em vias" OR "bloqueou as ruas" OR "coron virus" OR "corona voucher" OR "coronaa virus" OR "coronga vairus" OR "corono virus" OR coronovirus OR "corrona virus" OR "Curona Vairus" OR "curona virus" OR "Diwali In April" OR Ecoronavirusescuador OR "en primera linea") lang:en has:geo place_country:US

("essencial service" OR "essencial services" OR "essential service" OR "essential services" OR "face shield" OR "face shields" OR Frontline OR "hand sanitisers" OR "health worker" OR "health workers" OR homeschooling OR hydroxychloroquine OR "India for 21" OR "India Hum Honge Kamyab" OR "linha de frente" OR mascara OR mascarilla OR mascarillas OR "mascarillas desechables" OR "n95 mask" OR "no mask" OR "personal protective equipment" OR PPE OR "primera línea de la salud" OR quarentenou OR "Quarona Virus" OR remdesivir OR "servicio esencial" OR "servicios esenciales" OR "serviços essenciais" OR Shankh OR "trabajador sanitario" OR "Trabajadora sanitaria" OR "trabajadores de la salud" OR "trabalhador essencial" OR "trabalhadores essenciais" OR #LockdownLife OR #LockdownExtended OR #lockdowneffect OR #mortonaocompra OR #isolamentoparcial OR "isolamento parcial" OR #lockdownparcial OR #mortonaovota OR "passaportes de imunidade" OR "imunidade de rebanho" OR #UnitedAgainstCoronavirus OR #CovidISSNAF OR "testes em humanos" OR "human trials" OR gripezinha OR "the virus" OR "china virus" OR chinavirus OR "wuhan virus" OR wuhanvirus) lang:en has:geo place_country:US
